# A Randomized-Clinical Trial of Two Ambient Artificial Intelligence Scribes: Measuring Documentation Efficiency and Physician Burnout

**DOI:** 10.1101/2025.07.10.25331333

**Authors:** Paul J. Lukac, William Turner, Sitaram Vangala, Aaron T. Chin, Joshua Khalili, Ya-Chen Tina Shih, Catherine Sarkisian, Eric M. Cheng, John N. Mafi

**Affiliations:** Department of Pediatrics, David Geffen School of Medicine; UCLA Health Information Technology, UCLA Health, University of California, Los Angeles, Los Angeles, CA, United States; Division of General Internal Medicine and Health Services Research, David Geffen School of Medicine, University of California, Los Angeles, Los Angeles, CA, United States; Department of Pediatrics, Division of Immunology, Allergy and Rheumatology, David Geffen School of Medicine; UCLA Health Information Technology, UCLA Health, University of California, Los Angeles, Los Angeles, CA, United States; Department of Medicine, David Geffen School of Medicine, University of California, Los Angeles, Los Angeles, CA, United States; Department of Radiation Oncology and Department of Medicine, David Geffen School of Medicine; Department of Health Policy and Management, Fielding School of Public Health; Program in Cancer Health Economics Research, Jonsson Comprehensive Cancer Center, University of California, Los Angeles, Los Angeles, CA, United States; Division of General Internal Medicine and Health Services Research, David Geffen School of Medicine, University of California, Los Angeles, and Veterans Administration Greater Los Angeles Geriatrics Research Education and Clinical Center, Los Angeles, CA, United States; Department of Neurology, David Geffen School of Medicine; UCLA Health Information Technology, UCLA Health, University of California, Los Angeles, Los Angeles, CA, United States; Division of General Internal Medicine and Health Services Research, David Geffen School of Medicine, University of California, Los Angeles, Los Angeles, CA, United States; RAND Corporation, Santa Monica, CA, United States

**Author notes:** **Corresponding Author:** Contact information: 10833 Le Conte Ave, MDCC 22-432, Los Angeles, CA 90095.

## Abstract

**Importance:** Ambient artificial intelligence (AI) scribes record patient encounters and generate visit notes almost instantaneously, representing a promising solution to documentation burden and associated physician burnout. Despite swift and widespread adoption of AI scribes, their impacts have not been examined in randomized-clinical trials.

**Objective:** To test the effectiveness of two AI scribes in reducing time spent writing notes and associated burnout in a randomized-clinical trial.

**Design:** Parallel three-arm pragmatic randomized-clinical trial where physicians were assigned 1:1:1 via covariate-constrained randomization (balancing on time-in-note, baseline burnout score, and clinic days /week) to either one of two AI scribe applications—Microsoft DAX or Nabla—or a usual-care control group from 11/4/2024-1/3/2025.

**Setting:** A large academic health system in California.

**Participants:** 313 outpatient physicians were recruited based on leadership referrals and department-wide emails. 238 participants representing 14 specialties qualified.

**Intervention:** Intervention-arm physicians gained access to an AI scribe for two months.

**Main Outcomes and Measures:** The primary outcome was change from baseline log writing time-in-note. Secondary outcomes measured by surveys included Mini-Z 2.0, 4-item physician task load (TL), and Professional Fulfillment Index-Work Exhaustion (PFI-WE) scores to evaluate aspects of burnout, work environment, and stress, as well as targeted questions addressing safety and accuracy.

**Results:** DAX was used in 33.5% of 24,696 visits; Nabla was used in 29.5% of 23,653 visits. Nabla users experienced a 9.5% [95% CI:-17.2%,-1.8%] (p=.02) decrease in time-in-note versus the control group and a 7.8% [-15.5%,-0.1%] (p=.05) decrease versus DAX users, while DAX users exhibited no significant change versus control (-1.7% [-9.4%,+5.9%]; p=.66). Total Mini-Z, scaled 10-50 with higher scores indicating improvement, increased with users of any scribe (+2.76 [+1.41,+4.10]; p<.001). Reductions in TL (scale 0-400, TL=-35.8 [-63.7,-7.9]; p=.01) and work exhaustion (scale 0-4, PFI-WE=-0.27 [-0.48,-0.07]; p=.01) were seen with users of any scribe. One Grade 1 (mild) adverse event was reported, while clinically-significant inaccuracies were noted “occasionally” on 5-point Likert questions (DAX 2.7 [2.4-3.0] vs. Nabla 2.8 [2.6-3.0]; p=.68).

**Conclusion and Relevance:** Use of Nabla reduced time-in-note, while use of any scribe led to modest improvements in physician burnout, work exhaustion, and task load. Performance was remarkably similar across two distinct vendor platforms, and occasional inaccuracies observed in either scribe require ongoing physician vigilance.

**Trial Registration:** ClinicalTrials.gov Identifier: <u>NCT06792890</u>

## Introduction

Burnout afflicts nearly half of U.S. physicians, rising to endemic status and fueling a workforce exodus that amplifies an already critical physician shortage.^1–5^ Pernicious symptoms of exhaustion, depersonalization, and diminished sense of personal accomplishment have far-reaching consequences. The effects manifest across the healthcare ecosystem, jeopardizing care access, doubling risk of patient safety events, incurring billions of dollars in undue costs, and endangering physician well-being.^6–9^ While causes of burnout are complex and multi-factorial, electronic health records (EHR) and related documentation burden are frequently cited culprits.^10–14^ This excessive EHR charting forces physicians to spend as much as two hours documenting for every hour of direct care.^15,16^ A systematic review detailing EHR characteristics associated with burnout identified “insufficient time for documentation” as a top contributor.^17^

To address this problem, human scribes, both in person and virtual, have been deployed with some success, though these options present challenges around cost and accessibility, particularly among less well-resourced specialties.^18–21^ Digital scribes, which incorporate artificial intelligence (AI) to generate drafts that still require human editing by vendor staff, have shown variable impact, with little to no efficiency gain or burnout improvement, but generally positive user reception.^22–24^ Fully autonomous ambient AI scribes, which leverage large language models (LLMs), have been received with great enthusiasm by both industry and providers and are less expensive than human scribes, increasing scalability and the potential for widespread adoption.^25–28^ At present, over 50 ambient AI companies offer serviceable products, though few studies have rigorously evaluated their impact.^2,29–31^ Two small, nonrandomized pilot studies of DAX reported decreases in documentation time, EHR time, task load (TL), and aspects of burnout.^32–34^ However a larger, nonrandomized study using DAX showed no significant changes in financial and EHR use metrics, while discordantly, survey respondents in the intervention group subjectively reported decreased EHR time.^35,36^ Separately, in a large Nabla pilot, researchers reported small reductions in note time and favorable physician reception.^37^

Despite the many questions surrounding the use of AI in healthcare as well as the novel challenges posed by generative AI, there are a dearth of randomized-clinical trials evaluating its effectiveness and safety in real-world settings.^29,38–40^ In this trial evaluating two AI scribes, we aimed to investigate effects on documentation time and physician psychometrics, as well as AI scribe usability, accuracy, and safety. Additionally, with many options available to health systems, we sought to compare two leading vendors to inform future investment in this nascent technology.

## Methods

### Study Design

Enrolled physicians (N=238) were randomized 1:1:1 to either of two intervention arms, one provisioned Microsoft DAX^©^ and one provisioned Nabla^©^, or to a contemporaneous control group. To achieve cohort balance, covariate-constrained randomization was performed on physician baseline time-in-note (an Epic Systems, Inc^©^ [Verona, WI] Signal metric), a single item burnout score,^41^ and the number of self-reported clinic days/week. A mandatory pre-study survey was sent in September 2024, and a post-study survey was sent on January 4^th^, 2025. The trial was implemented during 11/4/2024-1/3/2025.

The study protocol was developed in accordance with Standard Protocol Items: Recommendations for Interventional Trials-Artificial Intelligence (SPIRIT-AI) and registered on ClinicalTrials.gov (<u>NCT06792890</u>).^42^ Study results reported here follow Consolidated Standards of Reporting Trials-Artificial Intelligence (CONSORT-AI).^43^ The UCLA Institutional Review Board (IRB-24-5425) determined that the study did not constitute human subjects research. Institutional urgency to rapidly test the tools during a brief contractual trial period precluded its timely pre-registration. The finalized trial protocol was submitted December 9, 2024, and published on January 27, 2025.

### Setting and Participants

Outpatient physicians were recruited via department-wide emails and nominations from department leaders. Physicians were required to hold at least one half-day of clinic/week, and those in intervention arms who previously used human scribes were obligated to forego this assistance during the study. Participants were instructed to use the AI scribe at English-only visits due to lack of internal validation of translation capabilities. **Figure 1** summarizes the trial recruitment and population.

**Figure 1.**
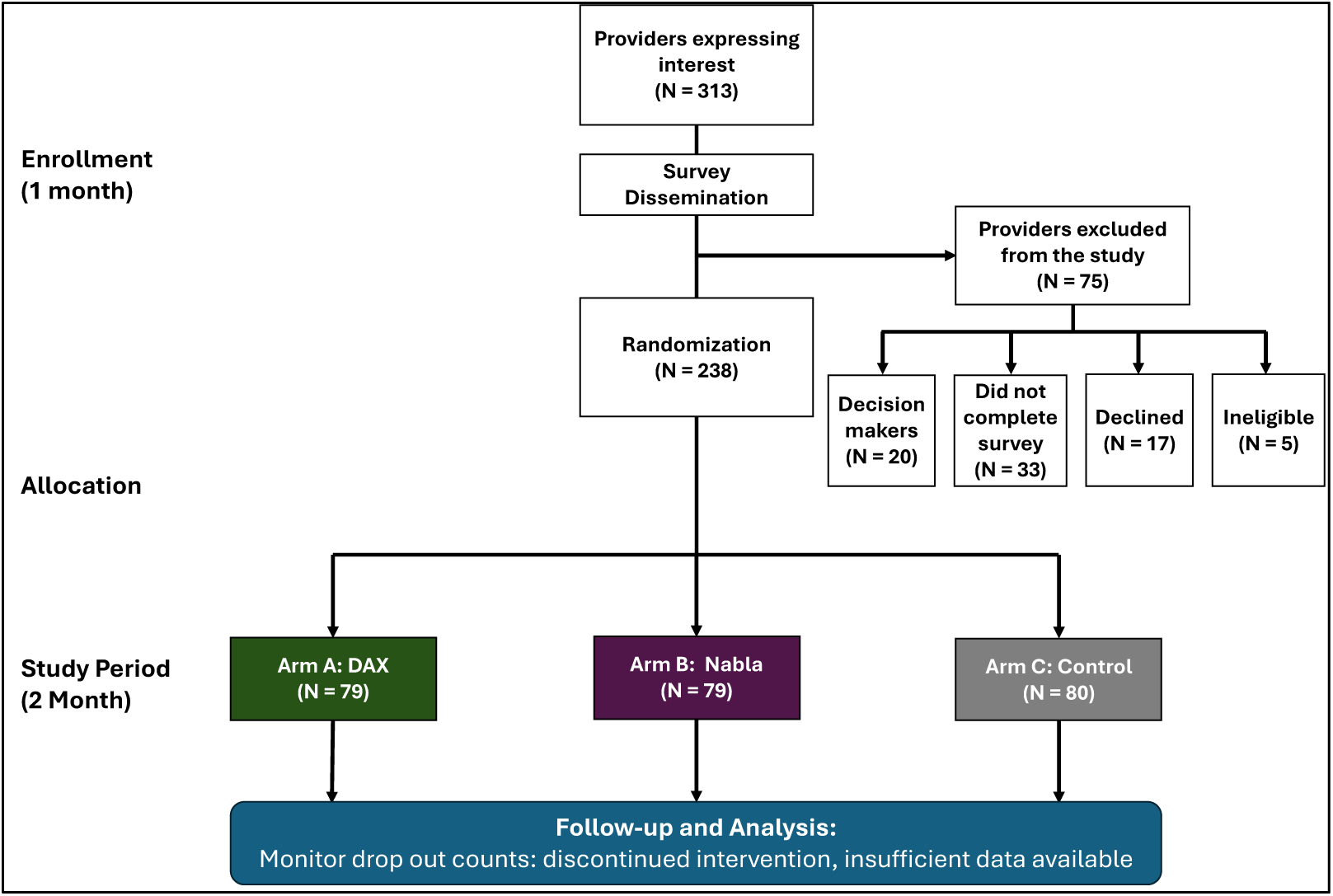
CONSORT-AI Flow Diagram.

### Interventions

DAX v2.0 and Nabla v1.5 were used. Both products were integrated into the EHR, allowing AI scribe-produced text to populate Epic’s native note types. Participants received one hour of application-specific training virtually, delivered by physician informaticists and vendor representatives, 1-2 weeks prior to study onset. Training was standardized for length, vendor participation, and format of internally generated educational material. Two weeks after study onset, vendors delivered a second presentation highlighting advanced features and customization. Ongoing support included an internal messaging channel and email listserv as well as vendor assistance. Participants were allowed to discontinue use at any time.

In compliance with California state law requiring two-party consent for audio recording, all physicians were instructed to obtain verbal consent from all relevant parties and document it in their note.

### Sample Characteristics

The pre-study survey, distributed via Qualtrics, characterized physician demographic and clinical time data.

### Primary Outcome

The pre-specified primary outcome was documentation time, specifically the amount of time spent writing a single note. For baseline comparison, time-in-note data were collected for the six months preceding study onset, while study data were collected from 11/4/24-1/3/25. We allowed one month “lead time” for participants to reach proficiency and thus only compared the second intervention month (12/2/24 to 1/3/25) to baseline.

### Secondary Outcomes

The pre- and post-study surveys included the following validated survey instruments (**Table 1**): Mini-Z 2.0 to assess burnout, work environment and pace, and EHR stress (10-50 scale, lower scores=worse burnout, more stressful workplace);^2,44,45^ 4-item physician task load, to assess cognitive load related to stress from EHR documentation (0-400 scale, lower scores=less cognitive load);^46,47^ and Professional Fulfillment Index–Work Exhaustion (PFI-WE 0-4 scale, lower scores=less exhaustion).^48,49^ The post-study survey addressed other topics via 5-point Likert questions, such as usability, occurrence of inaccuracies and biases, and perceived risks to patient safety.

**Table 1.** Survey Psychometric Descriptions.

| Survey (psychometric) | Format | Scoring (Range) | Interpretation |
| --- | --- | --- | --- |
| Mini-Z 2.0 (*burnout, satisfaction, stress*) | 10 questions divided into 2 subscales:<br>- Supportive Work Environment<br>- Work Pace & EMR Stress | - Total (10-50)<br>- Subscales (5-25)<br>- Single-item burnout question (1-5) | - Total score $\geq 40$ indicates a ‘joyful workplace’<br>- Subscale $\geq 20$ indicates ‘supportive workplace’ and/or ‘manageable work pace & EMR stress’<br>- Single-item $> 3$ indicates not burnt out |
| Nasa Task Load Index (*cognitive demand*) | 4 questions assessing cognitive demands of note-writing:<br>- Mental<br>- Physical<br>- Temporal<br>- Effort | - Total (0-400)<br>- Individual (0-100) | Total represents the sum of each question; lower scores indicate lower task load |
| Professional Fulfillment Index-Work Exhaustion (*work exhaustion*) | 4 questions:<br>- Dread about work<br>- Physical exhaustion<br>- Lack of enthusiasm<br>- Emotional exhaustion | - Total and individual: 5-point Likert (0-4) | Total represents the average of the 4 questions, where low values indicate less work exhaustion |

Abbreviations: EMR, Electronic Medical Records

An additional Signal metric, time in the EHR on unscheduled days, was collected, and baseline comparisons were conducted as with time-in-note.

### Sample Size Calculation

Study sample size was constrained by exclusion criteria and a contractual limitation stipulating a maximum of 100 concurrent users of each tool. A sample size of at least 79 providers per condition (the size of our smallest condition) provides 80% power to detect effect sizes as small as 0.50 standard deviations, assuming a two-sample t-test and a two-sided 0.025 significance level (2-fold Bonferroni correction for the comparison of each tool with the control condition).

Using pre-study data from a comparable timeframe, the estimated log-scale standard deviation of change for the time-in-note metric was approximately 0.31. This design thus provided sufficient power to detect a 15.5% relative improvement in the primary outcome. Given a baseline-year geometric mean time-in-note of approximately 4m 43s, this corresponds to an absolute difference of roughly 44 seconds. This effect size falls within the range of values reported in recent studies.^32,34,37^

### Statistical Analysis

Analyses of outcomes followed an intention-to-treat protocol. Time-in-note was log-transformed and compared to providers’ prior six-month baseline. A linear mixed-effects model, which included a study arm effect, a period effect (second month vs. first month), and the interaction of these terms, was utilized to model change in log-time-in-note. Linear contrasts were used to evaluate the effect of each tool vs. control in the second month (primary hypothesis). Survey-derived quantitative outcomes were analyzed using unadjusted linear regression models of the absolute change from baseline to post-intervention. Binary outcomes (e.g., dichotomized burnout) were analyzed using logistic regression models of post-intervention responses, adjusting for baseline response.

A post-hoc analysis was performed to evaluate the association between tool usage rate and change in (log) time-in-note and survey scores. Linear regression models evaluated the correlation between the proportion of encounters where the assigned tool was used and changes in outcomes.

A multiplicity-adjusted significance level of 0.025 was used for the primary analysis. For all other comparisons, we used a 0.05 significance level. Results were collected on 1/15/2025 and all analyses were performed during 1/16/2025-5/19/2025 using R v. 4.4.2 (https://www.r-project.org/).

## Results

### Study Participants

238 physicians representing 14 specialties enrolled in the trial (**Table 2**). Females were overrepresented (60.5%) and nearly half of providers (46.6%) reported to be 35-44 years old.

**Table 2.**
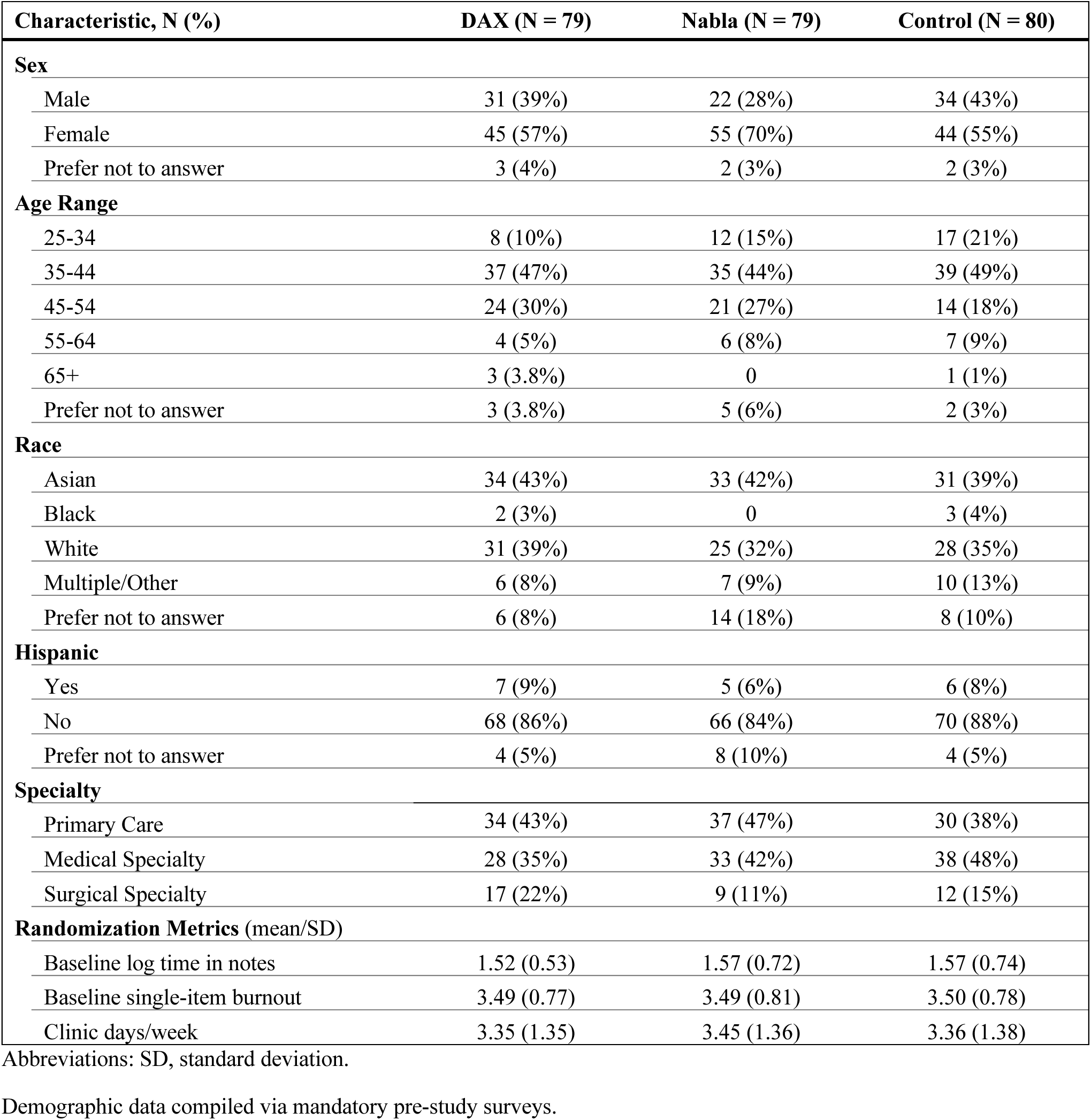
Study Physician Demographics.

DAX was used at 8,271 of 24,696 (33.5%) of patient visits, while Nabla was used at 6,981 of 23,653 (29.5%) visits. Control group encounters totaled 24,020. Approximately 15% of treatment-group physicians never used either tool.

### Primary Outcome

In our mixed model analysis, time-in-note declined by an estimated 18s (from 4m22s to 4m4s) in the control arm, 23s (from 4m29s to 4m6s) in the DAX arm, and 41s (from 4m30s to 3m49s) in the Nabla arm. The reduction in the Nabla arm was significantly larger than in the control arm (-9.5% [95%CI:-17.2%,-1.8%]; p=.02), while the reduction in the DAX arm did not significantly differ from control (-1.7% [-9.4%,+5.9%]; p=.66) (**Table 3**). The difference between these Nabla and DAX effects was itself statistically significant (-7.8% [-15.5%,-0.1%]; p=.05). Higher usage rate correlated with more time saved for either scribe, and this effect was more pronounced for Nabla users (r=-0.41) (**Figure 2A**).

**Figure 2.**
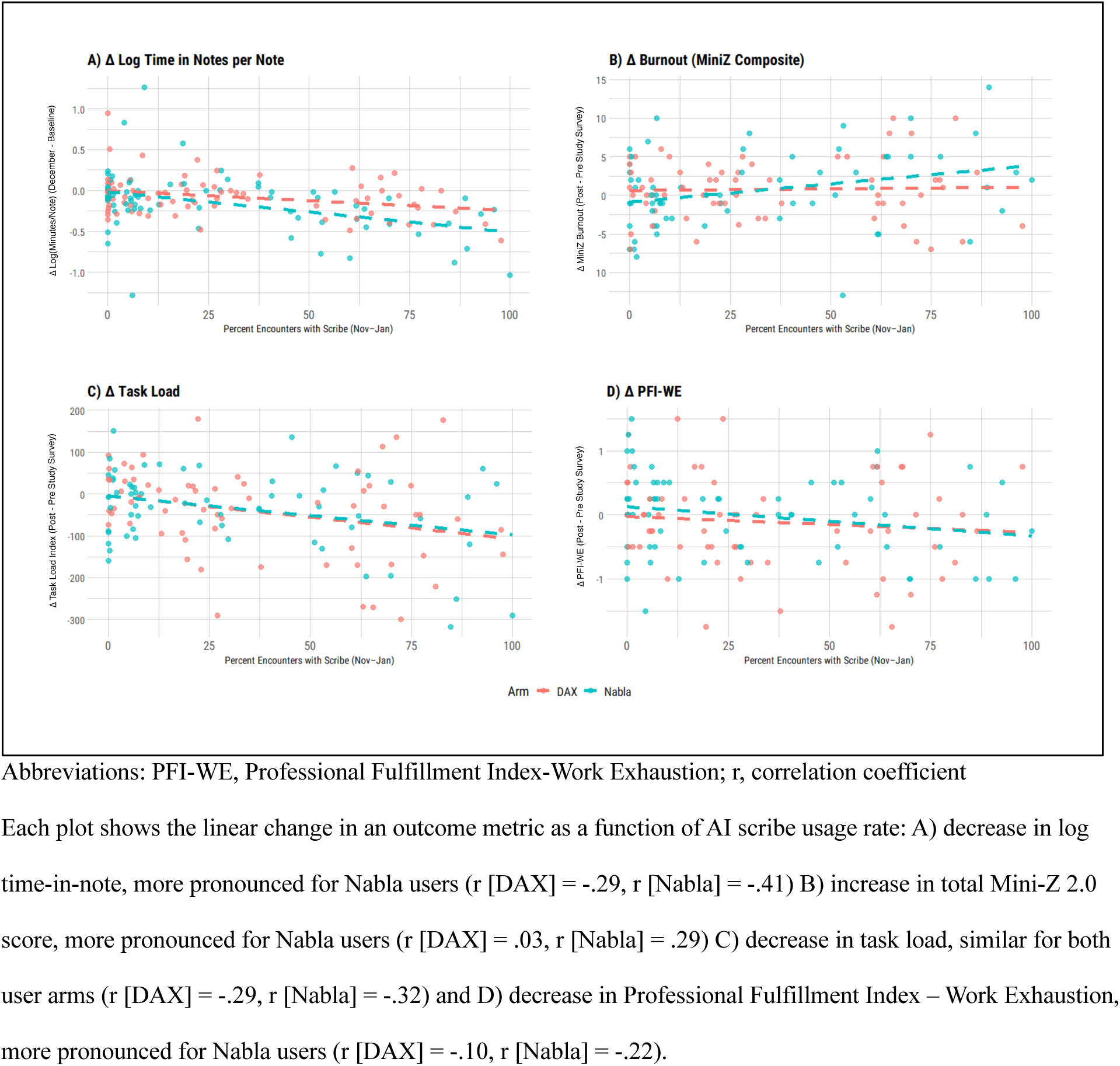
Individual Outcomes by Usage Rate.

**Table 3.** Psychometric Outcomes from Survey Assessments and Documentation Efficiency Metrics.

| Characteristic | DAX v Control |  |  | Nabla v Control |  |  | DAX v Nabla |  |  | Any v Control |  |  |
| --- | --- | --- | --- | --- | --- | --- | --- | --- | --- | --- | --- | --- |
|  | Difference | 95% CI | p-value | Difference | 95% CI | p-value | Difference | 95% CI | p-value | Difference | 95% CI | p-value |
| **Mini-Z 2.0** |  |  |  |  |  |  |  |  |  |  |  |  |
| Supportive Work Environment | 1.51 | [0.55, 2.47] | .002 | 1.55 | [0.59, 2.51] | .002 | 0.04 | [-0.90, 0.98] | .93 | 1.53 | [0.69, 2.37] | <.001 |
| Work Pace and EMR Stress | 1.41 | [0.38, 2.43] | .007 | 1.20 | [0.18, 2.23] | .02 | -0.20 | [-1.21, 0.80] | .69 | 1.30 | [0.41, 2.20] | .004 |
| Composite | 2.83 | [1.28, 4.37] | <.001 | 2.69 | [1.14, 4.23] | .001 | -0.14 | [-1.66, 1.38] | .86 | 2.76 | [1.41, 4.10] | <.001 |
| **PFI-WE** | -0.32 | [-0.55, -0.08] | .009 | -0.23 | [-0.46, 0.01] | .06 | 0.09 | [-0.14, 0.32] | .45 | -0.27 | [-0.48, -0.07] | .01 |
| **Task Load** | -39.89 | [-71.88, -7.90] | .02 | -31.69 | [-63.79, 0.42] | .05 | 8.20 | [-23.27, 39.68] | .61 | -35.79 | [-63.71, -7.87] | .01 |
| **Efficiency Metrics** |  |  |  |  |  |  |  |  |  |  |  |  |
| Time in Notes per Note | -0.02 | [-0.09, 0.06] | .66 | -0.10 | [-0.17, -0.02] | .02 | -0.08 | [-0.16, -0.00] | .05 | -0.06 | [-0.12, 0.01] | .10 |
| Time on Unscheduled Days | 0.14 | [-0.05, 0.33] | .14 | 0.06 | [-0.13, 0.26] | .53 | -0.08 | [-0.27, 0.11] | .42 | 0.10 | [-0.06, 0.27] | .22 |
|  | *OR* | *95% CI* |  | *OR* | *95% CI* |  | *OR* | *95% CI* |  | *OR* | *95% CI* |  |
| **Binary Mini-Z 2.0** |  |  |  |  |  |  |  |  |  |  |  |  |
| Single Item Burnout | 0.78 | [0.40, 1.54] | .48 | 0.67 | [0.34, 1.34] | .26 | 0.86 | [0.43, 1.71] | .67 | 0.73 | [0.40, 1.31] | .29 |
| Supportive Work Environment | 1.87 | [0.83, 4.23] | .14 | 1.84 | [0.82, 4.14] | .14 | 0.99 | [0.45, 2.15] | .97 | 1.86 | [0.91, 3.79] | .09 |
| Work Pace and EMR Stress | 1.90 | [0.17, 21.54] | .60 | 1.87 | [0.17, 21.19] | .61 | 0.98 | [0.13, 7.21] | .99 | 1.89 | [0.21, 17.25] | .57 |
| Composite | 1.64 | [0.38, 7.14] | .51 | 1.50 | [0.35, 6.45] | .59 | 0.91 | [0.24, 3.51] | .89 | 1.57 | [0.43, 5.76] | .50 |

Abbreviations: PFI-WE, Professional Fulfillment Index-Work Exhaustion; OR, odds ratio; CI, confidence interval

### Secondary Outcomes

The post-study survey was completed by 61/80 (76%) control group physicians and 65/79 (82%) physicians in each intervention arm. Total Mini-Z scores (scale 10-50) improved for users of any scribe vs. control (+2.76 [+1.41,+4.10]; p<.001), while TL scores (scale 0-400) and PFE-WE (scale 0-4) decreased for users of any scribe vs. control (TL=-35.8 [-63.7,-7.9]; p=.01; PFI-WE=-0.27 [-0.48,-0.07]; p=.01) **(Table 3**). Notably, there was no significant difference between the two scribes on any psychometric. As with time-in-note, higher usage rate correlated with greater improvement in Mini-Z and PFI-WE scores for Nabla users (Mini-Z: r=0.29; PFI-WE: r=-0.22) vs. DAX users (Mini-Z: r=0.03; PFI-WE: r=-0.10) (**Figure 2B,D**).

Inaccuracies were noted “occasionally” (DAX 2.7 [2.4-3.0] vs. Nabla 2.8 [2.6-3.0]; p=.68) and bias “rarely” (DAX 1.6 [1.4-1.8] vs. Nabla 1.7 [1.5-1.9]; p=.38) (**Table 4**). Respondents who noted “inaccuracies” and “bias” most frequently described omissions (n=12); structural concerns (e.g., undesired formatting, over-simplified language, or too much detail) (n=11); pronoun resolution errors (misgendering) (n=8); and affirmation/negation detection errors (n=5) (**Table 5**).

**Table 4.** Accuracy, Bias, and Usability.

| Question | DAX (N=65)<br>Mean (SD) | Nabla (N=65)<br>Mean (SD) | p-value |
| --- | --- | --- | --- |
| *Inaccuracies & Bias<sup>a</sup>* |  |  |  |
| In the last two months, how often did an ambient scribe-generated note contain at least one clinically significant inaccuracy (e.g., hallucination, omission, or addition)? | 2.7 (1.1) | 2.8 (1.0) | .68 |
| In the last two months, how often did an ambient scribe-generated note contain bias (e.g., unfair or skewed perspectives)? | 1.6 (0.9) | 1.7 (0.9) | .38 |
| *Usability<sup>b</sup>* |  |  |  |
| I used the tool as often as I could. | 3.8 (1.4) | 3.6 (1.4) | .45 |
| The tool was easy to learn. | 4.2 (1.1) | 4.1 (0.9) | .54 |
| The tool was easy to use. | 4.2 (1.1) | 3.9 (1.0) | .21 |
| The tool decreased the time I typically spend working on notes in clinic. | 3.7 (1.3) | 3.5 (1.2) | .33 |
| The tool decreased the time I typically spend in the EHR outside of work hours. | 3.6 (1.4) | 3.4 (1.2) | .34 |
| The tool decreased time it typically takes me to close patient encounters. | 3.8 (1.3) | 3.4 (1.3) | .15 |
| The tool is suited for the documentation needs of my specialty. | 3.6 (1.3) | 3.2 (1.3) | .10 |
| I could see myself using the tool in the future. | 4.1 (1.1) | 3.8 (1.3) | .19 |
| The tool generated notes at least as good as my own. | 3.4 (1.4) | 2.9 (1.3) | .06 |
| The tool allowed me to engage more with my patients. | 4.2 (1.0) | 3.8 (1.1) | .04 |
| Patients were generally okay with the use of the tool. | 4.4 (0.9) | 4.4 (0.9) | .77 |

Abbreviations: SD, standard deviation
<sup>a</sup> Response options: Never (1); Rarely (2); Occasionally (3); Frequently (4); Almost always (5)
<sup>b</sup> Response options: Strongly disagree (1); Somewhat disagree (2); Neutral (3); Somewhat agree (4); Strongly agree (5)

**Table 5.** Reports and Categorization of Inaccuracy and Bias.

| Category | Example | Vendor (n) |  |
| --- | --- | --- | --- |
|  |  | DAX (25) | Nabla (24) |
| ***Omission*** | “Omitted some pertinent information or at least information I would have liked to have for “smaller” problems. Had to either type myself or ask it to add in information and regenerate the note.” | 4 | 8 |
| ***Structure/Content*** | “As a specialist, there were frequently healthcare maintenance problems listed in the assessment and plan that were not relevant. Often too much detail as well. But overall it was helpful!” | 7 | 4 |
| ***Pronoun Resolution*** | “Gender was inaccurately identified.” | 8 | 0 |
| ***Affirmation/Negation Detection*** | “Somehow the scribe must have misheard the patient, but it reported “increase” rather than “decrease” for a lab result the patient mentioned. | 1 | 4 |
| ***Speech Recognition*** | “Misunderstood “not drinking alcohol” as “now drinking alcohol. Patient caught the error but was not angry, just wanted it corrected.” | 2 | 2 |

One adverse patient safety event was reported. Five physician co-authors (P.J.L., J.N.M., C.S., E.C., A.C.) independently deemed the patient safety event, described as “extensive patient counseling was not included in the assessment/plan or the patient instructions,” to be Grade 1 (mild).^50^

Of all usability questions, physicians rated both scribes lowest in response to the question, “The tool generated notes at least as good as my own,” where the average rating was still “neutral” (DAX 3.4 [3.1-3.7] vs. Nabla 2.9 [2.6-3.2]; p=.06). Meanwhile, both scribes were rated highly on improving physicians’ ability to engage with patients (DAX 4.2 [4.0-4.4] vs. Nabla 3.8 [3.5-4.1]; p=.04), while patients were receptive to their use (4.4 [4.2-4.6]) (**Table 4**).

There were no significant differences in time spent in the EHR during unscheduled days.

## Discussion

In this first randomized-controlled trial of LLM-powered ambient AI scribes, we observed a modest reduction in time spent on documentation amongst Nabla users, and improvements in burnout, work exhaustion, and task load with use of any scribe, relative to control. Exploratory analyses revealed a dose-response effect, wherein physicians who used an AI scribe at a higher percentage of encounters experienced greater benefit. Most physicians did not report inaccuracy and bias, found the AI scribes easy to learn and use, and felt the technology allowed them to better engage with their patients, results which may reflect additional mechanisms for reduced burnout aside from documentation time savings. For one of the most anticipated and rapidly adopted technological innovations in healthcare since the HITECH Act incentivized EHRs, these rigorous empirical findings both confirm prior observational studies showing positive results^32–34,37^ and reveal remarkably similar performance and reception across two distinct vendor platforms.

Various strategies to reduce burnout, ranging from individual approaches like peer coaching and stress management to organizational changes such as work hour reductions, have shown some success.^6,51,52,53^ At the same time, studies show clear limitations with existing interventions, such as costs of 1:1 peer coaching,^54,55^ insufficient data on long-term effects, and no clear preferred modality.^52^ There is growing anticipation that generative AI, when deployed to reduce physicians’ administrative and documentation burden, may represent a more scalable solution.^56,57^ Two prominent early use cases of generative AI, AI scribes and GPT-produced draft responses to patient messages (Epic Systems), have suggested some relief, even in the absence of large efficiency gains.^32–34,49^ In our study, AI scribe users experienced modest improvement in total Mini-Z 2.0 scores. However, a lack of significant improvement on the *binary* Mini-Z single-item burnout question or the subscales, which have been validated against an historical benchmark, the Maslach Burnout Inventory (MBI),^44,58^ limits interpretation of this finding though is likely due to insufficient statistical power. PFI-WE, itself validated against the MBI emotional exhaustion subscale and thus an alternative burnout indicator,^48^ and TL, established as a key mediator of EHR-related burnout,^46^ each decreased for users of any scribe. Taken together, these findings suggest that cautious optimism for AI scribes and their impact on physician wellness is warranted. Moreover, given our findings highlighting a relationship between greater usage and better outcomes, determining the best approach to engage physicians and maximize user experience remains an open question, one that we plan on investigating in future studies.

A central challenge to adopting AI scribes lies in justifying investment in this costly technology to health system leaders. In a recent report from Peterson Health Technology Institute, health system leaders identified increasing the “number of patient encounters per period” and “accuracy or completeness of coding for billing purposes” as two financial metrics.^31^ The former, however, contradicts the single most clear benefit documented in this study and others – physician wellbeing – and risks exacerbating physician disillusionment and exodus. Prior research suggests that burnout costs U.S. health systems $7,600 per physician per year (2015 dollars),^7^ translating to $36.5 million if applied to the 4,800 physicians employed at UCLA. A 7% reduction, as reported for our total Mini-Z and PFI-WE scores, could result in meaningful savings each year from decreases in burnout-associated turnover and reduced clinical hours.

When our physicians reported on inaccuracies, they highlighted multiple manifestations ranging from omissions to pronoun errors, magnifying the ambiguity and nuance in the clinical output of LLMs. Notably, there is a wide gap between our ability to *deploy* generative AI in healthcare at scale and our ability to *validate* generative AI at scale. As noted by Bedi, et. al,^59^ this is partly attributable to a lack of standardized tasks and dimensions of evaluation; for example, with AI scribes, should we focus on accuracy and factuality or omissions and comprehensiveness?

Historically, quality assessment of EHR documentation has relied on manual human review with frameworks such as the Physician Documentation Quality Instrument (PDQI-9).^60^ Given that LLMs are non-deterministic and subject to versioning, which can significantly influence output over time, relying on frequent end user feedback for quality assurance is both unrealistic and may itself exacerbate task load.^61^ One potential solution would be for LLMs to augment or replace human evaluators, i.e., LLMs as quality control agents.^62^ As noted by Croxford, et. al., this too comes with pitfalls, such as the rapid evolution of LLMs outpacing our ability to validate the “LLM evaluators”; LLMs’ inherent reliance on and sensitivity to prompts; and the challenge in replicating a physician’s nuanced clinical judgement that is necessary to determine if generated content is correct and meaningful in context of a patient’s clinical course.^63^ To be sure, the medical profession must embrace AI education and promote widespread AI literacy – essential steps toward safely and effectively integrating these tools into clinical practice.

Lastly, our study highlights several unique strengths and methodological insights. For one, a randomized-clinical trial represents the ideal method to control for selection biases, which is particularly important for optional workflows, such as whether to use an ambient scribe at a visit. Secondly, unlike static interventions in pharmaceutical trials, these tools rapidly evolve, even over the course of a study measured in months. Thus, a short, contemporaneous study period was advantageous in minimizing bias from such product changes while also avoiding the pitfalls of offering each vendor technology sequentially, where one product could mature more than the other. Finally, we demonstrated that AI scribes can be interchanged relatively seamlessly, which may speak to the ease of integration and use of this emerging technology.

### Limitations

First, this study was conducted at a single health system and thus our findings may not be broadly applicable. Second, our mix of participants were majority female, which could reduce generalizability given that females represent 38.1% of all physicians.^64^ However, the inclusion of multiple specialties strengthens overall generalizability. Third, our trial was relatively short as we were limited by contract terms, which stipulated both the length and number of users. This could limit the degree of impact given the time it takes to gain dexterity. The brevity may have also disincentivized physicians from investing time in learning the tool well and customizing it to their liking, which could lead to an underestimation of positive results. Nevertheless, despite this limitation, we still saw statistically significant positive outcomes. Fourth, physicians had the ability to edit the AI-generated note text in both the AI scribe platforms and in Epic’s note-writing interface, and we learned midway through the trial that Epic’s Signal metrics do not account for time in platforms. Hence, reported time-savings for time-in-note may represent an overestimation. Notably, this under-recognized limitation affects all AI scribe studies reporting Epic’s Signal metrics. Fifth, for the duration of the study, Nabla was not directly integrated within Epic’s mobile application (Haiku), while DAX was. Thus, Nabla users had to “launch” an encounter from a desktop computer to prompt a push notification to their phone. This may represent an important user interface difference affecting physicians’ usability impressions.

Sixth, there is a possibility of a selection bias given that some participants were identified by department leaders, though randomization minimizes this risk. There is also potential non-response bias associated with the post-study survey, which could skew survey results positively or negatively. However, response rates were similarly high across arms.

## Conclusion

In this randomized-clinical trial evaluating two ambient AI scribes, we observed modest yet broadly positive impact on physician outcomes, namely time-in-note for users of Nabla, and burnout, work exhaustion, and task load for users of any scribe. Future long-term studies will be essential to validating these trends across multiple institutions, establishing a robust cost-benefit analysis, measuring patient experience, and identifying clinicians who will benefit most from this technology.

## Data Availability

All data produced in the present study are available upon reasonable request to the authors.

## Acknowledgements

This project was supported by the UCLA Department of Medicine and the UCLA Faculty Practice Group. Dr. Mafi was supported by an NIH/NIA Beeson Emerging Leaders in Aging Research Career Development Award (No. K76AG064392-01A1). Dr. Sarkisian was supported by National Institutes of Health/National Institute on Aging (NIH/NIA) Midcareer Award in Patient-Oriented Aging Research (1K24AG047899) and NIH/NCATS UCLA Clinical and Translational Science Institute (CTSI) (UL1TR001881 PI Dubinett). We thank the UCLA Health Information Technology team for their technical support during the study and the participating physicians for their time and commitment. We appreciate the work of Chad Wes Villaflores, UCLA Healthcare Value Analytics Solutions Senior Data Scientist, for leading the ClinicalTrials.gov submission, Artem Romanov for his contributions to the IRB submission, and Katelyn Nguyen for her administrative support.

